# SARS-CoV-2 Protein in Wastewater Mirrors COVID-19 Prevalence

**DOI:** 10.1101/2020.09.01.20185280

**Authors:** Nafisa Neault, Aiman Tariq Baig, Tyson E. Graber, Patrick M. D’Aoust, Elisabeth Mercier, Ilya Alexandrov, Daniel Crosby, Stephen Baird, Janice Mayne, Thomas Pounds, Malcolm MacKenzie, Daniel Figeys, Alex MacKenzie, Robert Delatolla

## Abstract

The COVID-19 pandemic has given rise to diverse approaches to track infections. The causative agent, SARS-CoV-2, is a fecally-shed RNA virus, and many groups have assayed wastewater for viral RNA fragments by quantitative reverse transcription polymerase chain reaction (qRT-PCR) as a proxy for COVID-19 prevalence in the community. Most groups report low levels of viral RNA that often skirt the method’s theoretical limits of detection and quantitation. Here, we demonstrate the presence of SARS-CoV-2 structural proteins in wastewater using traditional immunoblotting and quantitate them from wastewater solids using an immuno-linked PCR method called Multiplex Paired-antibody Amplified Detection (MPAD). MPAD demonstrated facile detection of SARS-CoV-2 proteins compared with SARS-CoV-2 RNA via qRT-PCR in wastewater. In this longitudinal study, we corrected for stochastic variability inherent to wastewater-based epidemiology using multiple fecal content protein biomarkers. These normalized SARS-CoV-2 protein data correlated well with public health metrics. Our method of assaying SARS-CoV-2 protein from wastewater represents a promising and sensitive epidemiological tool to assess prevalence of fecally-shed pathogens in the community.

## INTRODUCTION

The measurement of SARS-CoV-2 RNA in wastewater has rapidly emerged as a means of gauging COVID-19 prevalence. Despite its promise, the approach has been limited by the high amplification levels necessitated, with Ct levels in the high 30s often observed, approaching the technology’s limit of detection and often exceeding the limit of quantitation. This could be a result of low starting fecal concentrations^1^ compounded by innate RNA lability in the harsh environment of sewage matrices that is potentially exacerbated by seasonal warmth.^2,3^ Further, sample preparation and viral RNA extraction adds considerable effort and costs for sewage surveillance programs. Assay sensitivity and cost are key considerations for wastewater-based infection early warning and monitoring. Finally, the limited viral RNA qRT-PCR assay sensitivity in raw wastewaters may preclude accurate and precise measurement of SARS-CoV-2 at strategic points in sewersheds upstream of a water resource recovery facility (WRRF), such as at specific institutional, commercial or residential nodes.

In contrast, SARS-CoV-2 proteins are present in higher copy numbers than viral RNA per virion^4^ and viral proteins in general are more stable^5,6^ than viral RNAs.^7^ Given the existence of MPAD (<u>M</u>ultiplex <u>P</u>aired-antibody <u>A</u>mplified <u>D</u>etection) technology which marries antibody specificity to PCR amplification, we hypothesized SARS-CoV-2 protein assessed by this approach might provide a stronger and more stable wastewater signal of SARS-CoV-2 presence than that derived from viral RNA. Therefore, we measured SARS-CoV-2 virus structural proteins in the same April-June, 2020 time series of wastewater samples from two wastewater recovery facilities (WWRF) in Canada’s capital Ottawa (pop. 1.1M) and the contiguous city Gatineau (pop. 300,000) for which we recently reported SARS-CoV-2 viral RNA.^8^ Despite comparatively low COVID-19 prevalence (<10 new daily cases per /100,000 inhabitants)^8^, analysis of sewage samples using standard immuno-blotting showed detectable levels of SARS-CoV-2 proteins. Quantitation of SARS-CoV-2 proteins in the samples by MPAD resulted in a signal which was orders of magnitude greater than that generated by qRT-PCR of viral RNA, correlating well both with quantified SARS-CoV-2 RNA as well as public health metrics across the 10-week study period. We believe MPAD based SARS-CoV-2 protein quantitation represents a promising epidemiological tool with a sensitivity sufficiently superior to viral RNA measurement that, in addition to enabling early detection and population tracking of COVID-19 load, will also open the way to effective infection surveillance of specific facilities, schools and residences.

## RESULTS

### Sample Nomenclature

Three fractions of wastewater are sampled and assayed in our study. Two sample fractions are collected from WRRF influent immediately following grit removal. WRRF influent is first degritted and then settled for one hour, and the resulting solids-rich sedimented layer is collected and referred to as “influent solids”. The remaining liquid fraction (supernatant) of the settled sample is then sequentially filtered through 1.5 µm and 0.45 µm filters, and the resulting filtrate is referred to as the “influent filtrate”. The third wastewater fraction studied is the “primary sludge”, obtained from the primary clarifier sludge stream (primary clarification is the processing step that immediately follows grit removal, where primary settling of wastewater solids occurs). The influent solids and filtrate were further concentrated via polyethylene glycol (or PEG) precipitation or filtration. The primary clarified sludge samples were either extracted directly or following PEG precipitation.

In addition, western blot analysis of SARS-CoV-2 protein was also made further along in the WWRF processing flow (see Supplementary Table 1 for sample descriptions).

### SARS-CoV-2 viral proteins are detected in primary sludge by western blot analysis

Primary sludge and PEG precipitated influent fractions, collected from the contiguous cities of Ottawa and Gatineau in April through June 2020, were analysed for the presence of four SARS-CoV-2 structural proteins, N (nucleocapsid), M (membrane), S (spike), and E (envelope), by western blot. In general, viral protein was detected exclusively in primary sludge and PEG precipitated influent solids with protein levels in PEG precipitated influent filtrates below the limit of western blot detection. The most prominent protein signal was detected in primary sludge samples, for which RIPA, urea and triton protein extraction buffers (see Methods) were used.

Nucleocapsid (N) protein was detected as a smear around 100kDa by two different anti-nucleocapsid (anti-N) antibodies from different vendors, directed at distinct N protein epitopes (see Supplementary Table 2), observed only in urea-extracted protein lysates from primary sludge samples from both municipalities at different dates (Figure 1). 100kDa corresponds to the anticipated size of N protein homodimers previously shown with size exclusion chromatography, and small-angle X-ray scattering by Zeng et al.^9^ In this regard, a band at 100kDa is also observed with longer exposure times in the purified protein lanes by both anti-N antibodies (Figure 1A). Signal extending to 50kDa, the size of monomeric N protein,^9^ was also observed in urea-extracted protein lysate from April 29 Gatineau primary sludge samples with both anti-N antibodies (Figure 1C). Higher molecular weight diffuse signal suggestive of possible higher order oligomerization was also observed in both primary sludge (Figure 1) and PEG precipitated influent solids (Supplementary Figure 1). In addition, the anti-N antibody (prosci) raised against the first 49 AA of the N protein detects a 20-25kDa band in triton-extracted samples from both municipalities (Figure 1), a possible reflection of nucleocapsid proteolysis previously described for Sars-CoV-1 N protein.^10^ Spike (S) protein, for which a ~180kDa band is anticipated,^11^ was harder to detect (Figure 2), although a signal of this size was observed in Gatineau May 14 primary sludge (Figure 2B) and to a lesser extent in Ottawa May 5 primary sludge (Figure 2A). In these samples, a lower MW band around 35kDa, which may be a truncated product of S protein was also detected. S protein is known to be glycosylated, and these post-translationally modified S protein or its subunits could explain the smeared signal seen on the western blots.^11,12^

**Figure 1:**
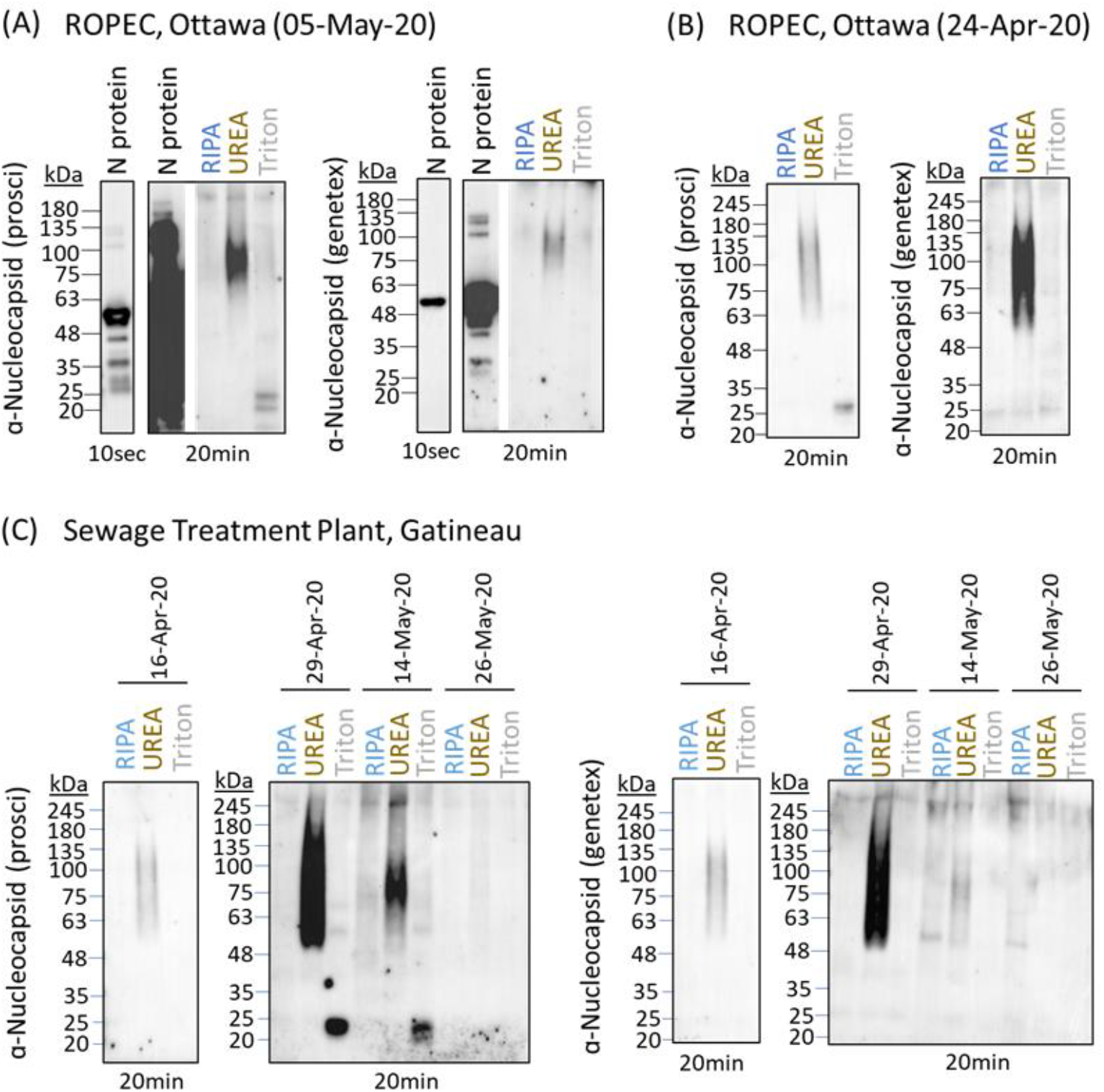
SARS-CoV-2 nucleocapsid (N) protein is detectable in primary sludge from wastewater in Ottawa and Gatineau by Western Blot. Protein was extracted from unconcentrated primary sludge samples from (A) Ottawa (May 5, 2020), (B) Ottawa (April 24, 2020) and (C) Gatineau (collected between April 16 and May 26), comparing RIPA, urea and triton X-100 lysis buffers. **Total protein lysate prior to processing by acetone-cleanup precipitation was resolved by 10% SDS-PAGE for the detection of N protein**.

Membrane protein is consistently detected at the expected size of 25kDa in both Ottawa (Figure 2A) and Gatineau primary sludge samples (Figure 3A) for RIPA, urea and triton protein extraction protocols. In contrast to the case of PEG precipitated influent solids, where a smaller ~18kDa band was observed (Supplementary Figure 1), bands at higher molecular weights (~35 and 46kDa, triton extraction, Figure 3A) are also present in some samples; whether these bands reflect non-specific detection by the anti-membrane antibody remains to be elucidated. All antibodies gave clear bands on the purified recombinant protein at molecular weights conforming to manufacturers specifications. Western blot analysis of the PEG precipitated influent solids fraction revealed lower amounts of protein with greater heterogeneity in signal consistent with harsher conditions and greater degree of protein degradation (Supplementary Figure 2).

**Figure 2:**
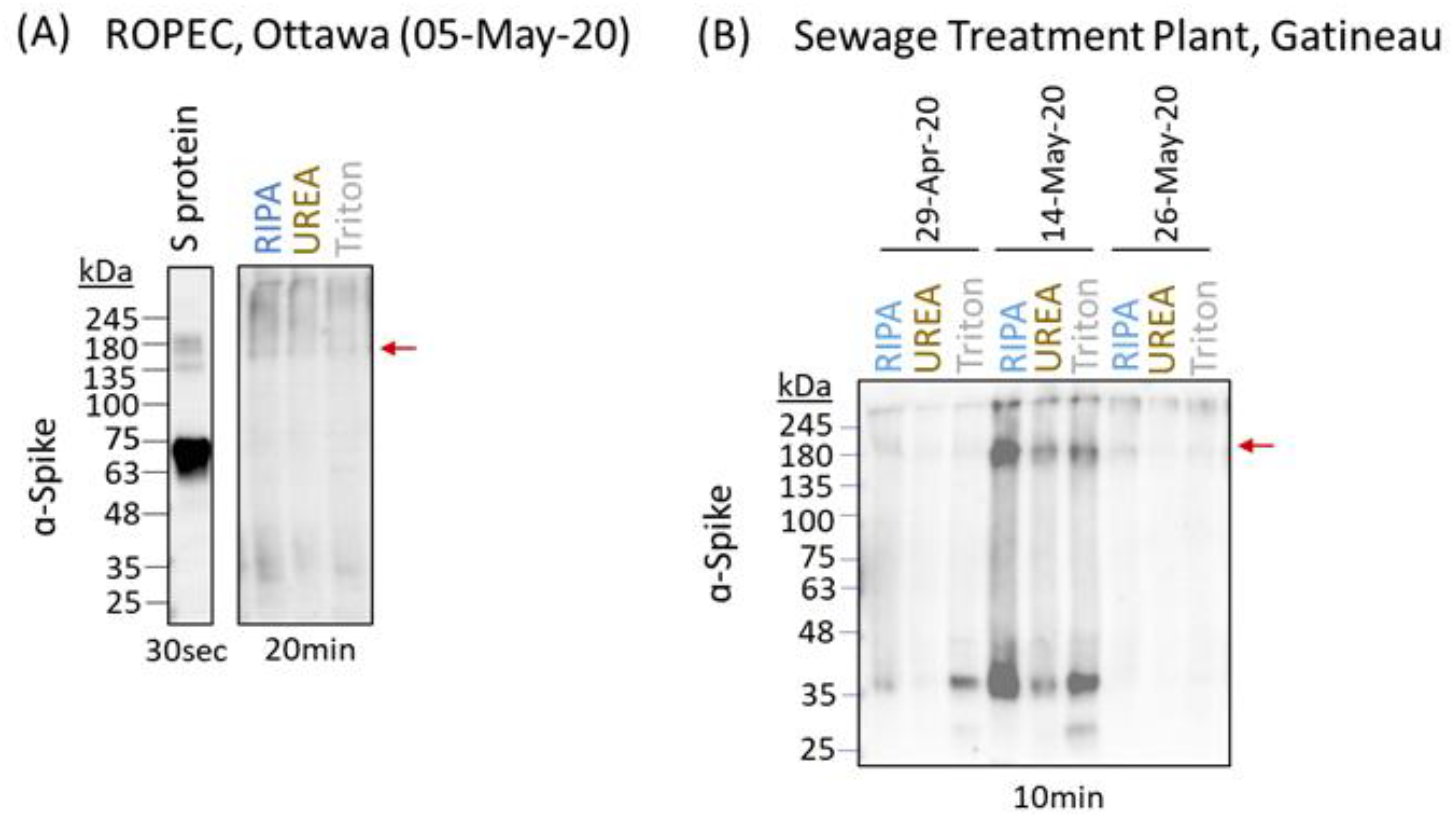
SARS-CoV-2 spike (S) protein is detectable in primary sludge from wastewater in Ottawa and Gatineau. Protein was extracted from unconcentrated primary sludge samples from (A) Ottawa (May 5, 2020) and (B) Gatineau (collected between April 29 and May 26, 2020), comparing RIPA, urea and triton X-100 lysis buffers. Protein lysates were subsequently cleaned up by acetone-precipitation, resuspended in RIPA lysis buffer with loading dye and resolved by 4-20% SDS PAGE for detection of S protein. Full-length S protein at 180kDa is indicated by red arrow.

### Protein Time Course

The 25kDa M protein band detected in primary sludge samples, as the most robust and consistently observed signal, was used to track community viral load in samples collected from Gatineau (Figure 3D). After normalizing the 25kDa signal to total protein, all three protein extraction methods showed a decline over the course of the Spring COVID-19 pandemic in Gatineau, paralleling that which is seen with viral RNA (Figure 3D) as well as being consistent with a downward trend of COVID-19 prevalence during the trailing phase of the pandemic in this community.

**Figure 3:**
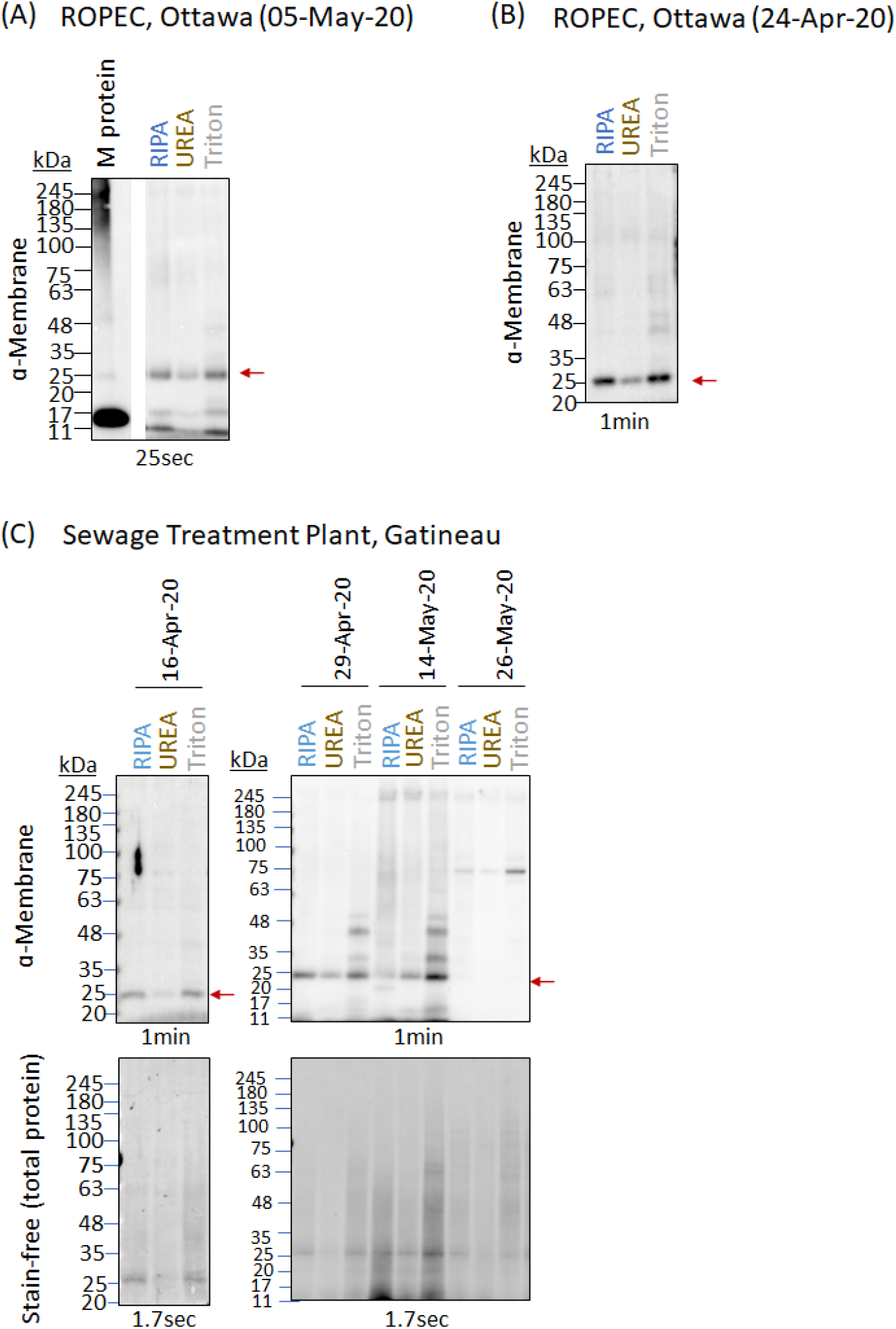
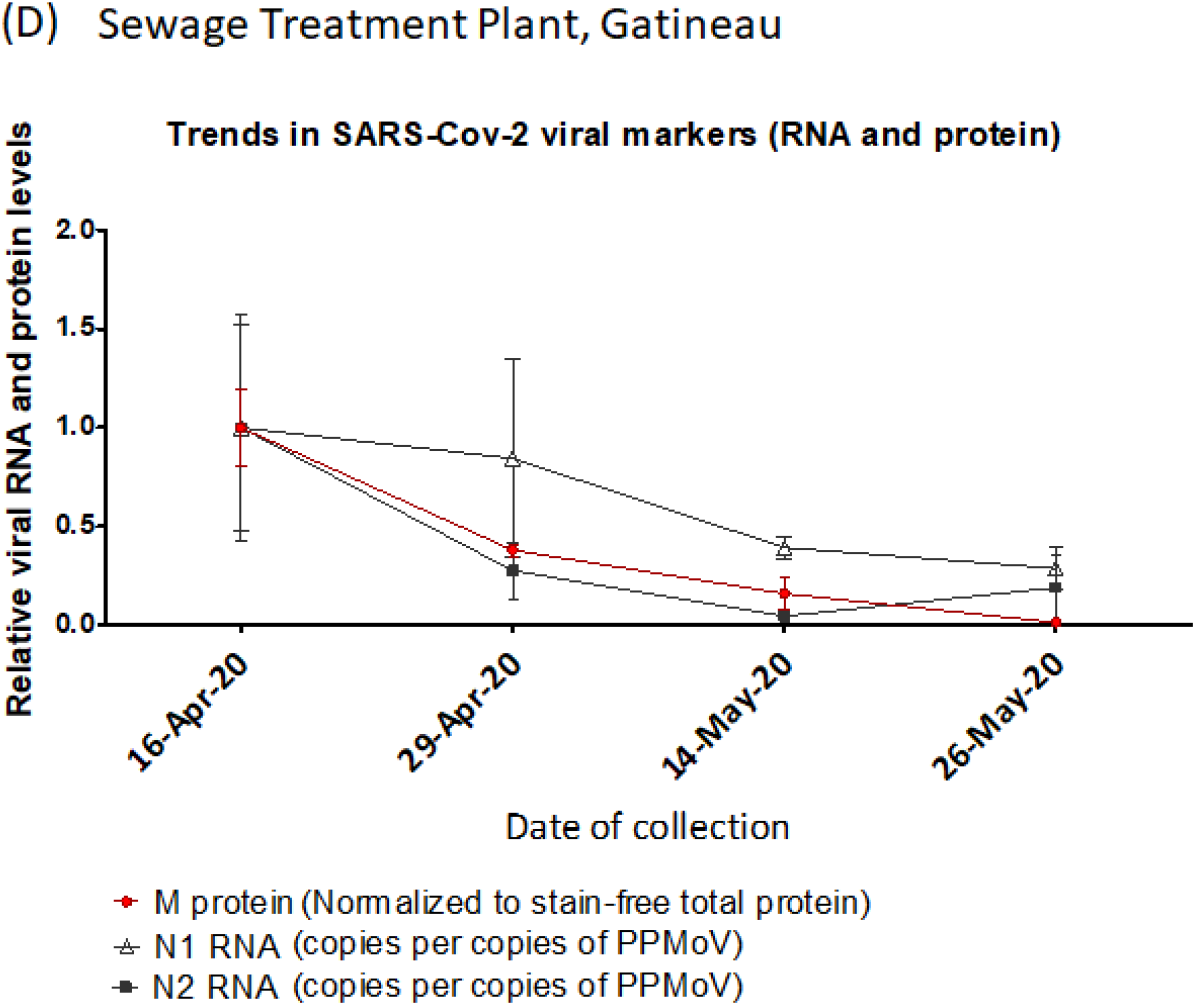
SARS-CoV-2 membrane (M) protein is detectable in primary sludge from wastewater in Ottawa and Gatineau and tracks RNA. Protein was extracted from unconcentrated primary sludge samples from (A) Ottawa (May 5, 2020), (B) Ottawa (April 24, 2020) and (C) Gatineau (collected between April 16 and May 26, 2020), comparing RIPA, urea and triton X-100 lysis buffers. Protein lysates were subsequently cleaned up by acetone-precipitation (see methods and materials), resuspended in RIPA lysis buffer with loading dye and subsequently resolved by **10% SDS-PAGE for the detection of M protein**. (D) Red circle: M protein signal at 25kDa (indicated by red arrow in Figure 3C) for the Gatineau time-series was quantified by densitometric analysis using ImageLab software, normalized to total protein as detected by the stain-free signal, and plotted as the average level of protein detected by RIPA, urea, and triton X-100 extraction methods. Y-values are protein levels relative to levels detected on April 16 (set to a value of 1.0). Wastewater sample collection dates are specified on the X-axis. Error bars represent standard deviation between different extraction methods (RIPA, UREA and Triton X-100). Black triangles and squares (From D’Aoust *et al*., 2020)^8^: quantification of viral RNA (N1 and N2, respectively) numbers from PEG precipitated primary sludge quantified by qPCR and normalized to PMMoV. Y-values are RNA levels relative to level on April 16 (set to a value of 1.0). Collection dates are specified on the X-axis.

### Quantitative wastewater protein analysis by MPAD

Following the western blot confirmation of viral protein presence in primary sludge, we next measured protein level using the MPAD platform, which is a multiplexed protein detection method which allows for amplification of the detection signal via quantitative PCR (qPCR). The use of this method is described in Meyer et. al. and Yaglom et. al.13,14

A custom MPAD SARS-CoV-2 protein profiling panel was developed, using different pairs of oligonucleotide-linked antibodies each, pair directed at an individual viral protein (see Supplementary Table 4). A positive signal occurs with the simultaneous binding of both antibodies to distinct epitopes of the same protein. The resulting proximity of the two oligos allows their simultaneous interaction by a bespoke linker oligonucleotide permitting subsequent amplification of any positive signal by qPCR. This design minimizes the non-specific binding seen in single antibody platforms which can preclude accurate multiplexing of protein detection.

In the initial analysis, SARS-CoV-2 N protein was measured in a 6 sample time series from April 24th to June 16th in primary sludge samples from Ottawa (Fig 4). The initial MPAD panel for N protein also measured three fecal content control proteins: PMMoV, CMV and human α-Tubulin. Normalization of the N protein signal to PMMoV alone (Figure 4A), or all three fecal markers (Figure 4B), both revealed similar ~50% increases from April 24 peaking on May 5 and 19 before a general decline.

**Figure 4:**
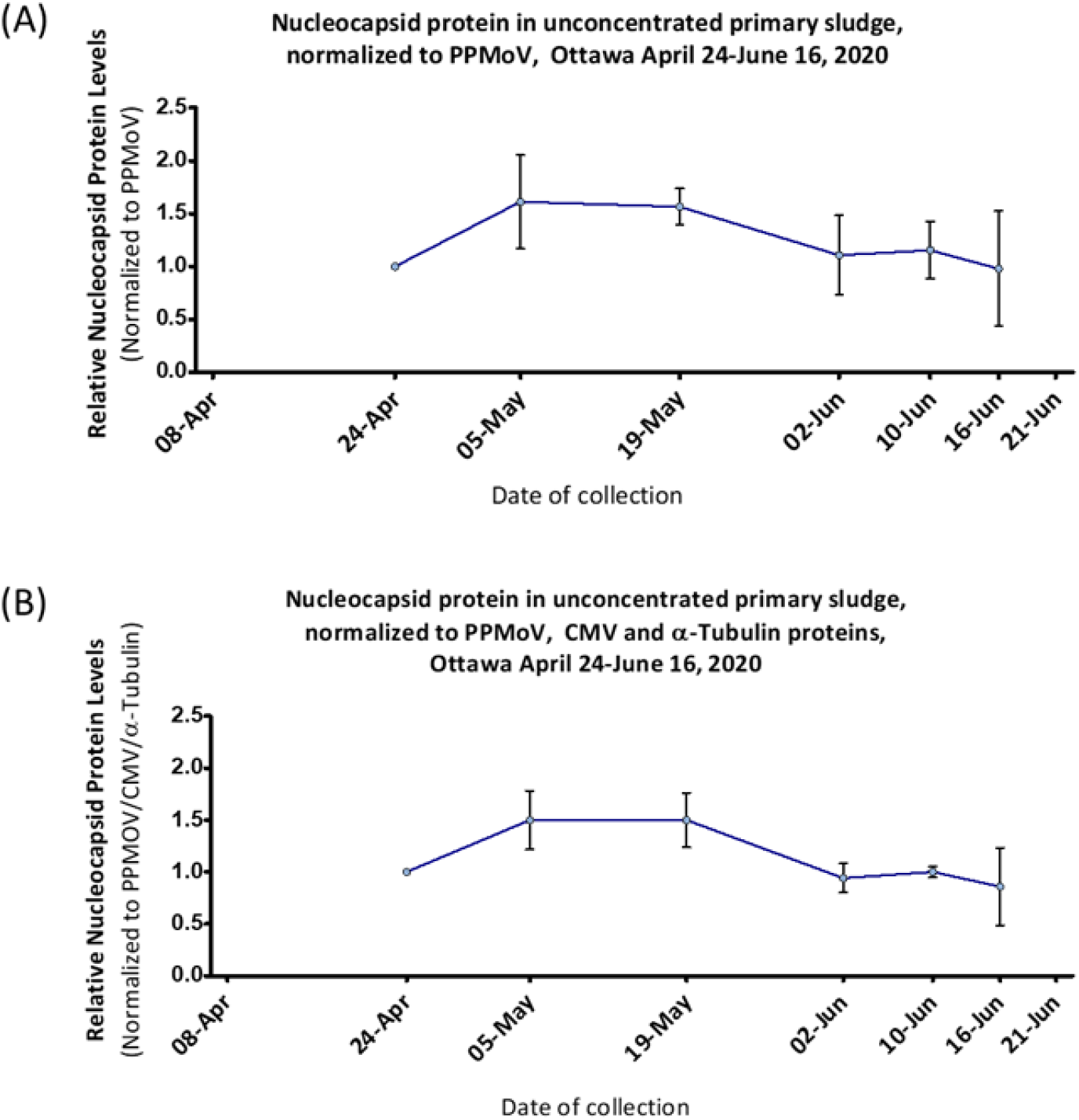
SARS-CoV-2 nucleocapsid (N) protein levels in Ottawa non-PEG-precipitated primary sludge determined by MPAD. N-protein from Ottawa non-PEG-precipitated primary sludge samples (collected between April 24 to June 16, 2020, X-axis, no samples were available for April 8 or June 21) was quantified by MPAD assay and normalized to (A) PMMoV alone or (B) PMMoV, CMV and α-Tubulin combined (geomean, see methods). Protein levels are plotted relative to April 24 reading (taken as 1). Error bars represent standard deviation between replicates.

Next, in order to assure specificity for detection of SARS-CoV-2 proteins, we used MPAD with an expanded panel to simultaneously measure three viral proteins, N, S and M, along with six fecal content control proteins in PEG precipitated “influent solids” samples drawn from the Ottawa WRRF during the study period (Fig 5). Inter-SARS-Cov-2 viral protein correlation between the three targets was good (Table 1, and see Methods) and the longitudinal analysis again showed a peak on May 5th followed by a slow decline (Fig 5), correlating broadly with N-protein in non-PEG-precipitated primary sludge (Fig 4). Together, these data strongly argue that 1) the structural proteins making up SARS-CoV-2 viral particles are detectable and measurable using MPAD in both unconcentrated primary sludge and PEG precipitated influent solids, and 2) these proteins co-vary in both sample types, suggestive of intact (although not necessarily infectious) virus particles. Finally, our normalization strategy combining signal (see Methods) from 6 different viral and human fecal content proteins successfully controls for a broad range of the variation inherent in wastewater based epidemiology monitoring, yielding a normalization superior to sewerage system metrics or single markers, maximizing accuracy and comparability between sample dates and sample fractions (Table 1, see Methods).

**Figure 5:**
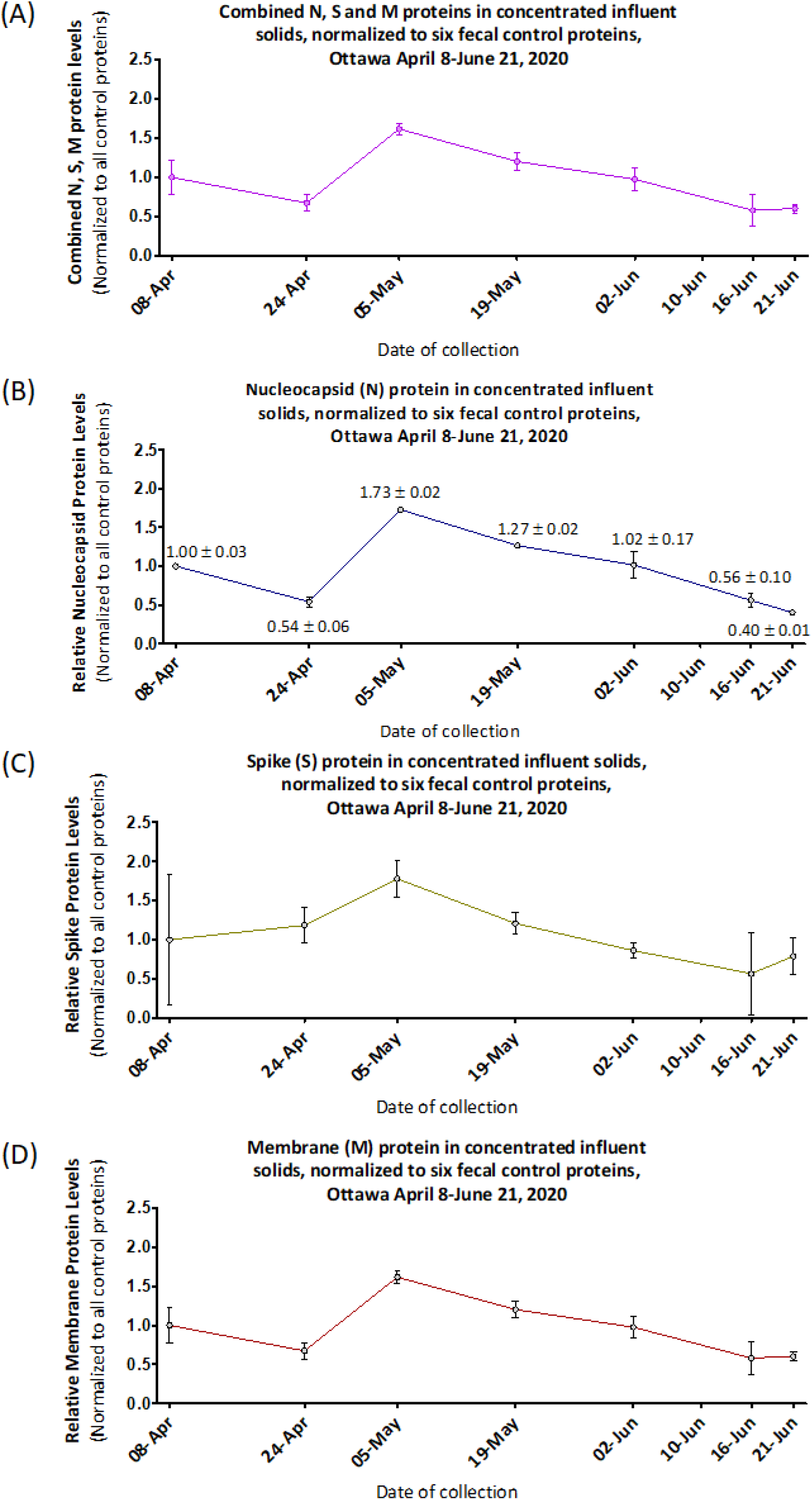
MPAD quantified SARS-CoV-2 structural proteins levels in Ottawa wastewater influent solids. N, S and M protein from PEG-concentrated influent solids from Ottawa (collected between April 8 and June 21, 2020, dates specified on the X-axis; no sample was analysed for June 10) quantified by MPAD assay normalized to the average of all six fecal control proteins: (A) N, S and M proteins combined, (B) nucleocapsid, (C) spike and (D) membrane protein. Data points are plotted as relative protein level compared to normalized viral protein in influent solids from April 8. Error bars represent standard deviation between replicates.

**Table 1:**
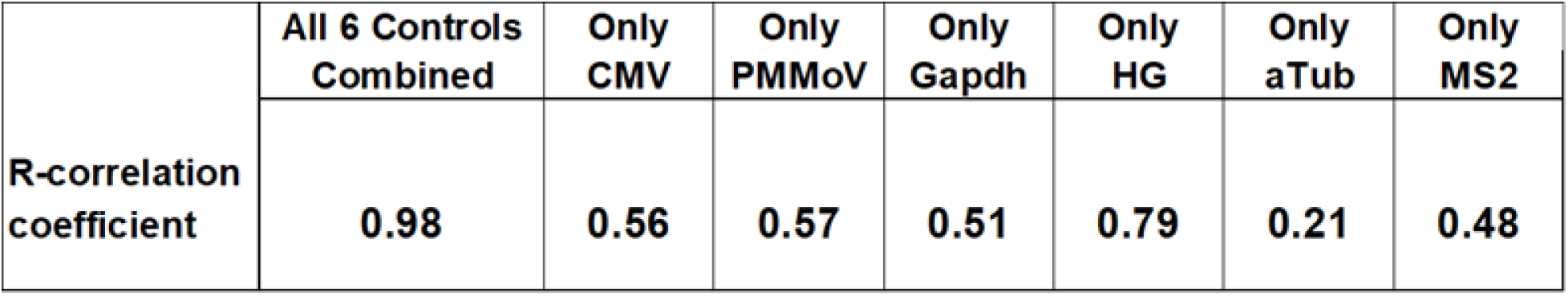
Inter-SARS-CoV-2 Protein Correlation for N, S and M proteins when normalized to single or combined control proteins quantified by MPAD. The multiple coefficients of correlation for N, S and M-proteins normalized to the geomean of all six of the fecal content control proteins combined (see methods), and to each individual control marker separately, were calculated for the seven sample dates (Figure 6). ^42^ Normalizing to combinations of fecal content control markers provides a superior basis for the viral protein measures compared to normalizing to any single fecal control protein.

|  | All 6 Controls Combined | Only CMV | Only PMMoV | Only Gapdh | Only HG | Only aTub | Only MS2 |
| --- | --- | --- | --- | --- | --- | --- | --- |
| **R-correlation coefficient** | **0.98** | **0.56** | **0.57** | **0.51** | **0.79** | **0.21** | **0.48** |

### Comparison of MPAD measured wastewater SARS Cov-2 protein to qRT-PCR quantification of SARS-CoV-2 RNA and community prevalence data; the Ottawa experience

MPAD based SARS-CoV-2 protein measurement was conducted on Ottawa primary sludge and PEG precipitated influent solids previously quantified for SARS-CoV-2 RNA by qRT-PCR.^8^ In this regard, viral RNA detection in the PEG precipitated influent solids was successful in four of these seven dates (average Ct 36.9), in contrast to successful protein quantitation of all samples by MPAD (average Ct 31), reflecting more protein than RNA in the samples, greater sensitivity of MPAD than qRT-PCR, or both. Moreover, the PEG precipitated influent solid viral RNA signal when detected did not correlate with viral RNA signal from PEG precipitated primary sludge, viral protein signal, or public health data (data not shown). Thus viral RNA signal in the PEG precipitated influent solids samples likely approximates the qRT-PCR limit of detection and does not accurately reflect wastewater viral titers, a finding consistent with our previous observation that N RNA sensitivity is higher in PEG precipitated primary sludge than in PEG precipitated influent solids.^8^

Given the viral protein detected in non-PEG-precipitated primary sludge by western blot (Figures 1–3), MPAD analysis was conducted on these samples, generating signal that correlated well with the MPAD trendline from PEG precipitated influent solids over the same time frame (Figures 4B and 5B, NB: Figure 4B starts only from April 24th). The trend for MPAD determined viral protein levels in PEG precipitated influent solids also paralleled that of a rolling 14-day sum of daily COVID hospital admissions, a metric selected as a proxy measure for total viral shedders in the community given low COVID prevalence and varying levels of daily individual testing (more detailed discussion in public health data section below) (Fig 6). PEG precipitated primary sludge viral RNA levels (both N1 and N2) measured by qRT-PCR are also shown for comparative purposes.

**Figure 6.**
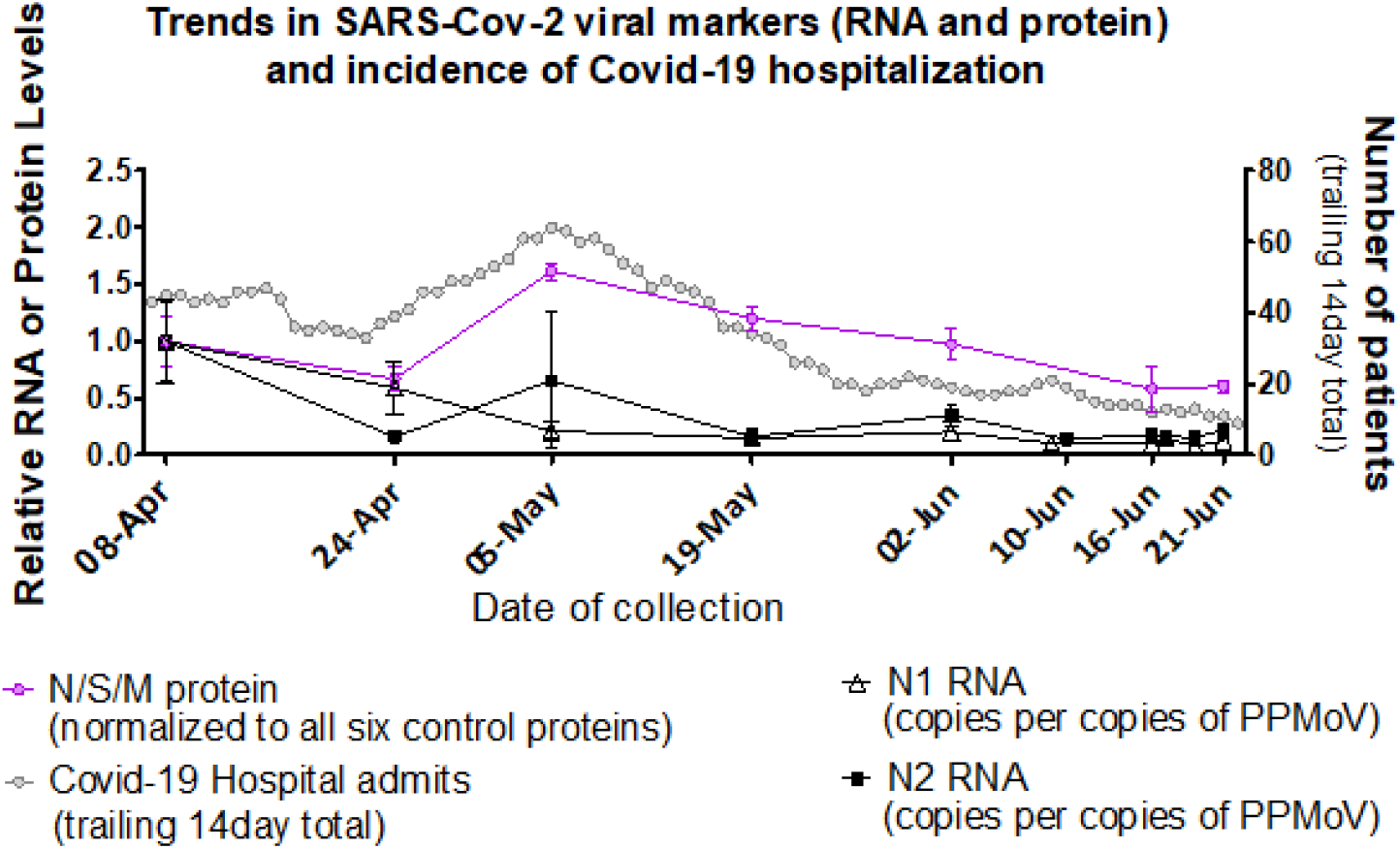
Levels of Ottawa wastewater SARS-CoV-2 structural proteins and RNA correspond with COVID-19 hospitalizations. Pink circles (from Figure 5): left Y-axis; MPAD determined Ottawa PEG precipitated influent solids N, S and M protein levels relative to April 8, 2020 (set to a value of 1.0), normalized to geomean of six fecal control proteins (see methods). Collection dates specified on the X-axis (no sample was analysed for June 10). Error bars represent standard deviation between replicates. Black triangles and squares (From D’Aoust *et al*., 2020)^8^: left Y-axis; RT-PCR generated Ottawa PEG precipitated primary sludge viral RNA (N1 hollow triangle and N2 solid square) relative to level on April 8 (set to a value of 1.0) normalized to PMMoV. Collection dates specified on the X-axis, including additional samples on June 17 and 19. Grey circles: right Y-axis, 14-day (estimated median duration of fecal viral shedding) trailing sum of daily hospital admissions, proxy for active viral shedders in the community.

### Comparison of MPAD measured wastewater SARS Cov-2 protein to qRT-PCR quantification of SARS-CoV-2 RNA and community prevalence data; an International Perspective

The many studies of SARS-CoV-2 viral RNA in municipal wastewater, despite coming from disparate regions and differing in approach, all share one thing: comparatively low RNA levels. In our own experience^8^ and the additional 7 manuscripts summarized in Table 2, the average Ct required for viral RNA detection was over 37 (vs. MPAD’s protein detection average Ct of 30-31). Categorizing the eight papers in Table 2 by sample fraction analyzed--primary sludge, influent solids, or influent filtrate, reveals greater viral RNA signal in the two solids fractions compared to influent filtrate, as would be expected for enveloped viruses.^15^ Also, predictably, the greater the degree of sample concentration and COVID-19 population prevalence the higher the viral RNA signal. Although with respect to prevalence data, as has been noted broadly, the low and inconsistent levels of COVID-19 testing in individuals, especially early in the pandemic, followed by an increase in testing levels over time in most locales make COVID-19 individual testing data an imperfect metric when assessing true community prevalence.^16^ Therefore, for calculations presented in Table 2, we have relied on cumulative cases as a serviceable proxy for prevalence, as it is less reliant on testing levels of individuals in specific periods. We also assume that areas that reported higher cumulative confirmed cases (per 100K population) likely also had higher levels of COVID-19 infection at the time of sewage sampling, leading to higher viral titers in wastewater.

**Table 2:**
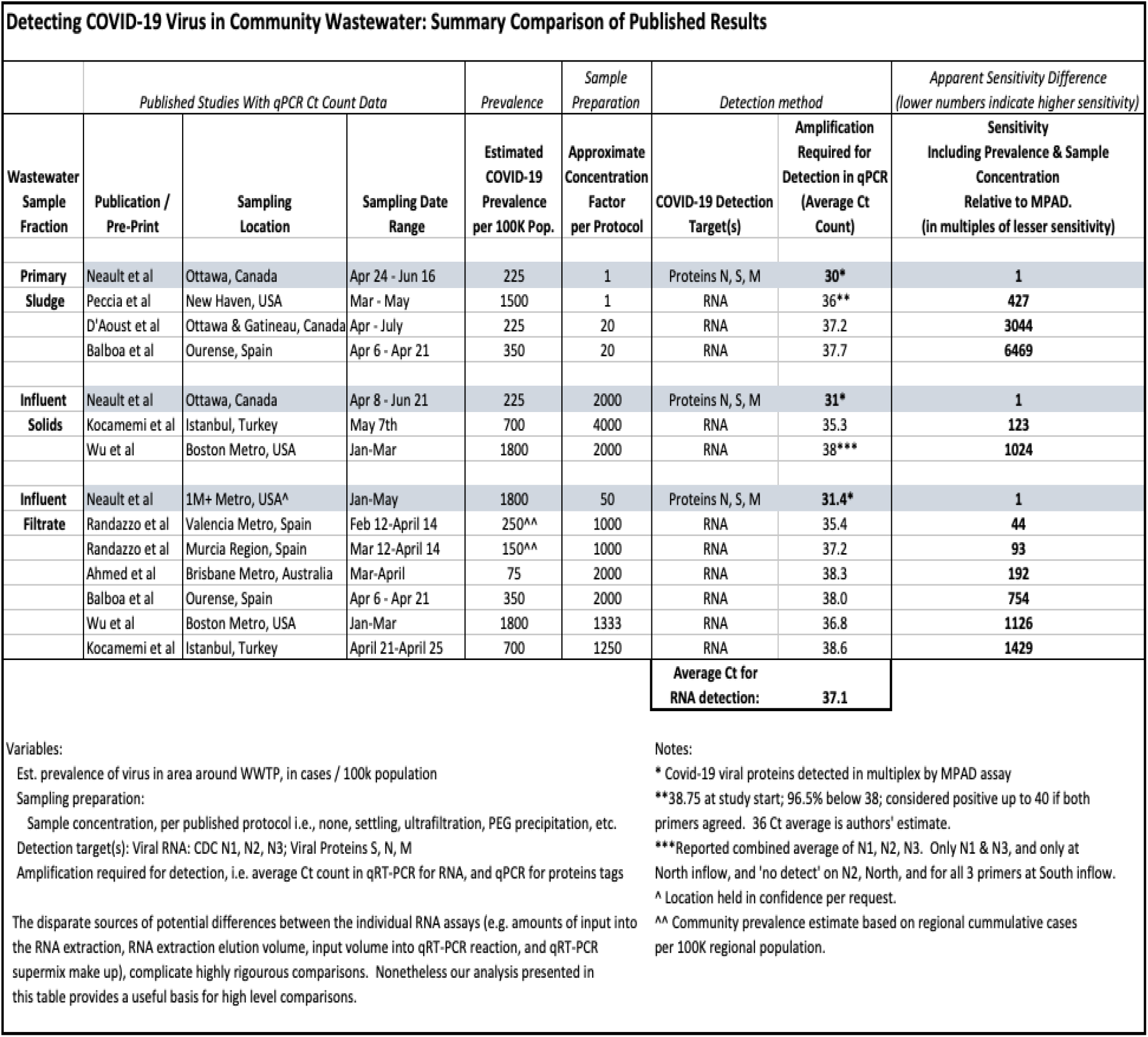
Published SARS-CoV-2 component levels in community wastewater and Covid prevalence. N.B. The low and inconsistent levels of individual COVID-19 testing, especially early in the pandemic, followed by increases in testing levels over time, make COVID-19 individual testing data an imperfect metric with which to assess true community prevalence.^16^ Therefore, we use cumulative community cases as a serviceable proxy for prevalence, as it is less reliant on testing levels of individuals in specific periods.^8,18,20,22–24,43–46^

MPAD measured viral protein in primary sludge generates a signal 64-208 fold (6 to 7.7 Cts) greater than the viral RNA signal determined by qRT-qPCR. For influent solids MPAD protein signal is 20X to 128X (4.3 to 7 Cts) greater than qRT-PCR viral RNA levels; and for influent filtrate MPAD protein signal is 16X to 147X (4 to 7.2 Cts) greater than qRT-PCR viral RNA levels.

These are the differences for unadjusted RNA and protein signals; however, the reports being compared in Table 2 are from cities ranging from low/medium (Ottawa) to high COVID-19 prevalence (New Haven, Boston, Istanbul) and there is no adjustment for disease prevalence; in addition there are differences in sample preparation protocols leading to different sample concentrations for each protocol. When adjusted for disease prevalence (see Table 2 legend) and degree of sample concentration, the difference between protein and RNA signal is even more marked, with protein readings ranging from over 400 to almost 6,500 times higher in primary sludge and approximately 100 to 1000 fold greater in influent solids. For the influent filtrate, for which time series samples from a US city were analyzed, the protein levels range from 40 to over 1400 fold greater than RNA signal (Table 2).

## DISCUSSION

The low wastewater SARS-CoV-2 RNA signal detected by our group and others may devolve from multiple factors, including low community COVID-19 prevalence, the exact sewage fraction sampled, the sample collection, handling and preparation approaches used, the degree of sample concentration, the presence of inhibitors, and, not least, the innate fragility of RNA in the harsh wastewater matrix. Moreover, the actual stool concentration of SARS-CoV-2 RNA itself is low (Cts in the high 30s) when compared with respiratory tract levels (Cts <30).^1^

Regardless of its source, issues with SARS-CoV-2 RNA detection represent a real challenge to the meaningful deployment of this technology, particularly in the sentinel surveillance of low prevalence populations, and for providing the earliest possible warning of initial infection outbreaks within a community. It is upon this context, we present here the detection of SARS-CoV-2 structural proteins in wastewater samples by immunoblot analysis and followed by its MPAD based quantification.

### SARS-CoV-2 Protein in Wastewater

Our western blot analyses revealed the presence of the four SARS-CoV-2 structural proteins in wastewater, a result which aligned well with Raman spectroscopy analysis showing persistence of spike protein throughout a series taken at different point of Wuhan wastewater treatment plant, in contrast to SARS-CoV-2 RNA which was found only in the most upstream samples.^17^

Membrane protein generates a strong signal at the expected molecular weight via western blotting, across all 3 extraction approaches (Figure 3). Moreover, M protein level trend-line from longitudinal samples detected by western blotting, and normalized by total protein, corresponded well with the viral RNA trend-line from those same samples (Figure 3D).

For nucleocapsid, the two different antibodies recognize the same protein band validates that the antibodies are binding specifically. Even within this fraction there was some heterogeneity however; the variable band sizes observed for nucleocapsid using the three different extraction buffers may devolve from differential solubility and stability of the various N protein species in the different systems. For example, the diffuse 100kDa bands correlating to putative dimerized N protein were uniquely observed in samples extracted by urea lysis buffer (Figure 1), which, unlike RIPA or triton X-100, is a chaotropic agent which can dissolve otherwise insoluble protein fractions. And while urea extraction gave the strongest nucleocapsid signal, RIPA and Triton yielded the strongest bands for the membrane protein.

### MPAD Sensitivity and Specificity

The MPAD platform (see Results and Methods) is a hybrid immunoassay technology allowing multiplex protein detection. The qPCR required for both MPAD and qRT-PCR methodologies allows direct comparisons of amplification cycle numbers, and thus protein to RNA signal comparison as well. As can be seen in Table 2, MPAD detects a protein signal at least one order of magnitude greater than RNA signal and sometimes 1000s of times greater. In regard to specificity, when conducting qRT-PCR viral RNA analysis, several genes are frequently targeted in order to increase assay specificity (e.g. SARS-CoV-2 N and E genes). In an analogous fashion, unlike other protein assays such as ELISA, the MPAD platform allows simultaneous analysis of multiple proteins in one sample; in the current study, to achieve optimal specificity, three viral proteins were assayed. Thus, specificity as one of the potential benefits of RNA detection is comparable with MPAD assay capability.

### Fecal Content Controls For Normalizing Viral Measures

Standardization of systems as variable as wastewater represents an important facet of generating accurate epidemiology data. In this regard, SARS-CoV-2 RNA wastewater studies range from reporting no normalization controls, to using WRRF system metrics as controls (e.g. daily system flow or mass flux), to sample process controls (e.g. extracted mass), to measuring surrogate organisms such as Pepper Mild Mottle Virus (PMMoV) known to be shed in human excreta.^8,16,18^–^24^ We recently presented normalization of viral RNA signal in PEG precipitated primary sludge using extracted mass, solids mass flux, and PMMoV showing PMMoV to be the most reliable of the three approaches,^8^ agreeing with other studies.^18,19^ Thus while the inherent variability (e.g. flow and wastewater characteristics) of a WRRF system contributes to wastewater viral RNA fluctuations, factors such as RNA stability within sewage matrices and other sampling related factors are also likely at play and the optimal marker for the fecal content should ideally reflect all of these factors. In this regard, while PMMoV-based normalization reduced the longitudinal variation in SARS-CoV-2 RNA signal,^8^ other studies have shown a combination of content markers to be superior to single marker normalization.^25,26^ Therefore, taking advantage of MPAD’s extensive multiplexing capacity, six fecal markers were added to the MPAD panel alongside the three SARS-CoV-2 proteins; PMMoV and MS2,^27,28^ cytomegalovirus (CMV, like SARS-CoV-2, an enveloped virus) known to be present in human stool^29^ and human α-tubulin, GAPDH and hemoglobin.^30^–^32^ To our knowledge, this is among the first wastewater studies to use the latter three human proteins as markers of fecal contribution to wastewater. N, S and M-proteins were each then normalized to the six fecal content control markers individually and to the combination of the 6 markers (Methods sections, Table 1 legend for details). The inter-marker correlation for N,S and M was then calculated for the two classes, i.e. levels normalized to individual controls versus normalized to aggregate controls (Table 1) showing that the combination of six control markers yields a substantially superior normalization, maximizing accuracy and comparability between sample dates and sample fractions. The robust specificity and normalization devolving from measuring all 9 (3 SAR-CoV-2, 6 fecal control markers) protein levels with this single, multiplex protein panel represents a significant advantage in any broad waste-water surveillance program.

### Ottawa Public Health Metrics for COVID-19 Infection

Overall, Ottawa experienced only modest levels of COVID-19 infection (active cases peaking at ~600, new daily cases of around 20, and never exceeding 60, general population >1M); the resulting relatively low viral load in wastewater was useful in benchmarking the sensitivity of novel viral detection methodologies. In terms of determining community prevalence, Ottawa tracks a variety of COVID-19 public health metrics^33^ including hospitalizations, daily COVID-19 tests, daily positive COVID-19 tests, and active cases. However, the latter three measures are clearly contingent upon the availability and targeting of testing; in this regard, Ottawa’s daily test numbers over the first four weeks experienced a 5 fold increase fueled in large measure by the opening of the test availability to the general public. Clearly then, these indices are unlikely to faithfully reflect true COVID-19 population prevalence. In contrast, hospitalizations for COVID-19 are based on comparatively invariant clinical assessment and ascertainment is high. Thus, as previously utilized by others,^34^ while only representing a ‘tip of the iceberg’ small percentage of all infected (given larger number of milder and even asymptomatic COVID-19 cases), we believe daily hospital admissions serve as a proxy for all viral shedders in a community and trends in these numbers should parallel viral protein measured in the wastewater (Fig 6).

Whether measuring wastewater SARS-CoV-2 protein or RNA to estimate population trends in COVID-19 prevalence, timing of viral stool shedding in relation to clinical activity becomes a central issue. Disparate periods of SARS-CoV-2 RNA fecal shedding have been proposed;^19,35^–^41^ taken together these analyses suggest an approximate median of 14 days of viral shedding. Thus, in using hospital admissions as a proxy for all viral shedders, these results suggest the daily admissions summed over the 14 days (the median expected viral shedding period upon hospital admittance) prior to a given date is indicated. In this fashion, we hope to include all current shedders on a given date who have been hospitalized. Figure 6 shows the resulting daily hospital admission trailing total for Ottawa over the wastewater sampling period, superimposed with the combined viral N, S and M proteins and RNA in wastewater. The two-week spacing of wastewater sampling to some degree complicates comparison to the hospital admission-based metric but good concordance is nonetheless seen.

Ultimately, determining to what extent wastewater viral protein can serve as a prevalence leading indicator will require a longer study with more frequent sampling over rising and falling community prevalence in order to assess robustly. However, it is encouraging to observe that the protein viral metrics in this study tracked well with this limited estimate of community viral load and with a sensitivity greatly exceeding that of viral RNA tracking.

## MATERIALS and METHODS

### Wastewater sample collection

Influent and primary sludge samples were collected from water resource recovery facilities located on the Ontario and Quebec banks of the Ottawa River in the National Capital Region of Canada serving over 1.3 million people. More specifically, samples were collected from Robert O. Pickard Environmental Centre (ROPEC) in Ottawa, ON and the sewage treatment plant in Gatineau, QC as previously described.^8^ Three types of samples were collected for this study: i. influent solids, ii. primary sludge, and iii. influent filtrate. In addition, influent filtrate samples were acquired from a large US city (that requested anonymity) and concentrated by ultrafiltration, for analysis by MPAD. Supplemental Table 1 describes the characteristics of the various wastewater samples assayed. These samples were used as-is (unconcentrated) or subjected to PEG precipitation (concentrated), as previously described.^8^

### Western Blots

#### Protein Extraction

Wastewater samples collected at various stages throughout the wastewater treatment facilities were processed in the lab within 24 hours of collection for influent solids and filtrate, and the primary sludge was initially stored at −80°C and thawed on ice prior to protein extraction. Protein was extracted from each sample using one of three lysis buffers in the presence of 1X Halt Protease and Phosphatase Inhibitor Cocktail (Thermo Scientific, Cat no. 1861281): urea buffer (5.25M urea, 1.5M thiourea, 3%CHAPS, 22.5mM Tris-HCl pH8.5); radioimmunoprecipitation assay buffer, RIPA (150mM NaCl, 1% Triton X-100, 0.5% Sodium deoxycholate, 0.1% Sodium dodecyl sulfate (SDS), 50mM Tris-HCl pH 7.4, 1mM EDTA); or triton X-100 buffer (1% triton x-100, 2mM phenylmethylsulfonyl fluoride (PMSF), 1X PBS pH7.4). Samples were lysed on ice for 20min with intermittent vortexing, then sonicated using the Bioruptor pico sonication device (Diagenode) at 4°C for 8 cycles (15sec on/60sec off) and cleared by centrifugation at 4°C at 23,000xg for 30min. Subsequently, the supernatant was transferred to new tubes for downstream work-up.

#### Acetone-precipitation and cleanup of protein lysate for western blots

Some extracted protein lysates were subjected to acetone-mediated precipitation to remove salts, lipids and other contaminants. Five volumes of chilled (to −20 °C) precipitation buffer (50% acetone, 50%ethanol, 0.1% acetic acid) were added to one volume of wastewater protein lysate, mixed by inversion and incubated at least overnight at −20°C to allow for protein precipitation. The precipitated protein was centrifuged at 4°C at 16,000xg for 25min, and the resulting protein pellets were subsequently washed three times in 0.5 ml ice-cold acetone by vortexing (5×15-30sec on max speed) and centrifuged at 4°C at 16,000xg for 10min after each wash. Finally, the pellets were air-dried with the tube cap open at room temperature for 10-20min and resuspended in one volume of RIPA buffer.

#### SDS-PAGE and Immunoblotting

Protein lysates were prepared for loading in a final 1X Laemmli sample buffer (BioRad, Cat No. 1610747) containing 2.5% 2-mercaptoethanol and heated at 95°C for 5 min; if the sample contained urea, it was incubated at room temperature only. For the detection of nucleocapsid and membrane SARS-CoV-2 proteins, approximately 15µl of protein lysate was resolved on a 1.5mm 10% SDS-PAGE gel using the 10% TGX stain-free fast-cast kit (BioRad, Cat no. 1610182). For the detection of spike and envelope SARS-CoV-2 proteins, 7.5µl of lysate was run on a 1mm gradient SDS-PAGE gel using pre-cast 4–20% Mini-PROTEAN TGX stain-free protein gels (BioRad #4568096). The corresponding positive controls shown in Supplementary Table 2 were also loaded and the gels were run at 100V. Post-run, the stain-free signal was activated by 45sec exposure under UV using the ChemiDoc imaging system (BioRad) and transferred to low fluorescence polyvinylidene fluoride (LF PVDF) membranes using the Trans-Blot Turbo RTA kits (BioRad, Cat no. 1704270 or 1704271) and the semi-dry TurboBlot turbo transfer system (BioRad). The primary antibodies in Supplementary Table 3 were used to detect the respective protein on the western blots; blots were incubated overnight at 4°C with primary antibody diluted in 1% milk in TBS-T (1X TBS, 0.05% Tween 20) and 0.1% sodium azide. Blots were subsequently washed 3×10min with TBS-T at room temperature, incubated with HRP-linked secondary antibodies which were diluted 1:2500 in 5% milk/TBS-T 1hr at room temperature – goat anti-mouse IgG (H+L) HRP conjugate (BioRad, Cat no. 1706516) or goat anti-rabbit IgG (H + L)-HRP conjugate (BioRad, Cat no. 1706515) – and washed again for 3×10min in TBS-T, 2×5min in TBS at room temperature. Chemiluminescence signal from antibody complexes was activated using Clarity ECL western blotting substrate (BioRad, Cat no. 1705060 or 1705061) and visualized using the camera-based ChemiDoc imaging system (BioRad). Quantification of stain-free total protein and target-protein chemiluminescent signals was performed by densitometric analysis using the ImageLab software (Bio-Rad).

### <u>M</u>ultiplex <u>P</u>aired-antibody <u>A</u>mplified <u>D</u>etection (MPAD) of SARS-CoV-2 viral proteins

In the current project, pairs of oligo-tagged antibodies were used in a custom MPAD protein profiling panel (ActivSignal, Natick, MA), against spike (S), nucleocapsid (N) and membrane (M) SARS-CoV-2 proteins, as well as antibodies against fecal content control proteins: i) PPMoV, ii) Bacteriophage MS2, iii) cytomegalovirus (CMV), iv) α-Tubulin, v) glyceraldehyde 3-phosphate dehydrogenase (GAPDH) and vi) hemoglobin--all combined in a single multiplex panel. The paired antibodies for each protein target were carefully selected and tested in combination. The antibodies used are outlined in Supplementary Table 4. After incubation of the sample with conjugated antibodies, the oligo barcodes from bound antibodies are collected and analyzed using the Fluidigm BioMark HD microfluidic qPCR platform (South San Francisco, CA).

#### MPAD assay data normalization

The limitations of normalizing with a single control gene or protein have been previously reported.^26^ The MPAD protein analysis panel consisted of six fecal content controls (proteins from PPMoV, Bacteriophage MS2, and cytomegalovirus (CMV), the human proteins α-tubulin, glyceraldehyde 3-phosphate dehydrogenase (GAPDH) and hemoglobin) as well as the three SARS-CoV-2 viral proteins (N, M and S protein). A normalization method combining the six fecal control proteins previously described for gene expression was used.^25^ Ct values are first converted to bioload using the 2^-(Ct) formula, followed by calculation of bioload geomean of all samples for each protein. The bioload for each protein is then divided by this geomean to determine the “geomean ratio”. Next, a ‘sample geomean’ was calculated for each sample across the geomean ratios (from step 2) of all the fecal control targets. This fecal control sample (or replicate) geomean was then divided into the bioloads for each of the viral proteins, to calculate the normalized value for that sample (or replicate) for that protein. As a final step, the average and standard deviation of replicates was taken for each sample.

A multiple coefficient of correlation between N, S and M-proteins was calculated for the seven sample dates normalized to all six of the fecal content control proteins combined, and to each individual control marker separately--using the approach outlined at <u>Multiple Correlation</u>.42

#### MPAD Negative Controls and Titration

Supplemental figure 3 shows an Ottawa PEG precipitated influent solids sample taken on April 8^th^ compared to and with signal substantially above: i. a PBS negative control; and, ii. a ‘no-linker’ negative control. Supplemental figure 4 shows Recombinant N protein samples diluted in PBS 3 times, with a dilution of 10X between the samples, analyzed by the MPAD assay. The titration shows linearity of the signal, and is above background signal in the PBS negative control and a ‘no-linker’ negative control.

## Data Availability

Data is available from the authors.

## References

1. Collivignarelli, M. C. et al. SARS-CoV-2 in sewer systems and connected facilities. Process Safety and Environmental Protection (2020) doi:10.1016/j.psep.2020.06.049.

2. Rosa, G. La, Bonadonna, L., Lucentini, L., Kenmoe, S. & Suffredini, E. Coronavirus in water environments: Occurrence, persistence and concentration methods - A scoping review. Water Res. 179, 1–11 (2020).

3. Hart, O. E. & Halden, R. U. Computational analysis of SARS-CoV-2/COVID-19 surveillance by wastewater-based epidemiology locally and globally: Feasibility, economy, opportunities and challenges. Sci. Total Environ. 730, 1–9 (2020).

4. Bar-On, Y. M., Flamholz, A., Phillips, R. & Milo, R. SARS-CoV-2 (COVID-19) by the numbers. Elife 172, 697–698 (2020).

5. Stone, N. P., Demo, G., Agnello, E. & Kelch, B. A. Principles for enhancing virus capsid capacity and stability from a thermophilic virus capsid structure. Nat. Commun. 10, 1–13 (2019).

6. Russell, C. J., Hu, M. & Okda, F. A. Influenza hemagglutinin protein stability, activation, and pandemic risk. Trends Mictobiology 26, 841–853 (2018).

7. Wada, M., Lokugamage, K. G., Nakagawa, K., Narayanan, K. & Makino, S. Interplay between coronavirus, a cytoplasmic RNA virus, and nonsense-mediated mRNA decay pathway. Proc. Natl. Acad. Sci. U. S. A. 115, E10157–E10166 (2018).

8. D’Aoust, P. M. et al. Quantitative analysis of SARS-CoV-2 RNA from wastewater solids in communities with low COVID-19 incidence and prevalence. *medRxiv* 1–40 (2020).

9. Zeng, W. et al. Biochemical characterization of SARS-CoV-2 nucleocapsid protein. Biochem. Biophys. Res. Commun. 527, 618–623 (2020).

10. Mark, J. et al. SARS coronavirus: Unusual lability of the nucleocapsid protein. Biochem. Biophys. Res. Commun. 377, 429–433 (2008).

11. Walls, A. C. et al. Structure, Function, and Antigenicity of the SARS-CoV-2 Spike Glycoprotein. Cell 181, 281-292.e6 (2020).

12. Fung, T. S. & Liu, D. X. Post-translational modifications of coronavirus proteins: Roles and function. Future Virol. 13, 405–430 (2018).

13. Meyer, R. D. et al. TMIGD1 acts as a tumor suppressor through regulation of p21Cip1/p27Kip1 in renal cancer. Oncotarget 9, 9672–9684 (2018).

14. Yaglom, J. A. et al. Cancer cell responses to Hsp70 inhibitor JG-98: Comparison with Hsp90 inhibitors and finding synergistic drug combinations. Nat. Sci. Reports 8, 1–12 (2018).

15. Ye, Y., Ellenberg, R. M., Graham, K. E. & Wigginton, K. R. Survivability, Partitioning, and Recovery of Enveloped Viruses in Untreated Municipal Wastewater. Environ. Sci. Technol. 50, 5077–5085 (2016).

16. Medema, G., Heijnen, L., Elsinga, G., Italiaander, R. & Brouwer, A. Presence of SARS-Coronavirus-2 RNA in Sewage and Correlation with Reported COVID-19 Prevalence in the Early Stage of the Epidemic in The Netherlands. Environ. Sci. Technol. Lett. 7, 511–516 (2020).

17. Zhang, D. et al. Ultra-fast and onsite interrogation of Severe Acute Respiratory Syndrome Coronavirus 2 (SARS-CoV-2) in environmental specimens via surface enhanced Raman scattering (SERS). medRxiv 86, 2020.05.02.20086876 (2020).

18. Wu, F. et al. SARS-CoV-2 titers in wastewater are higher than expected from clinically confirmed cases. medRxiv 21, 1–14 (2020).

19. Wu, F. et al. SARS-CoV-2 titers in wastewater foreshadow dynamics and clinical presentation of new COVID-19 cases. medRxiv 53, 1689–1699 (2013).

20. Ahmed, W. et al. First confirmed detection of SARS-CoV-2 in untreated wastewater in Australia: A proof of concept for the wastewater surveillance of COVID-19 in the community. Sci. Total Environ. 728, 138764 (2020).

21. Ahmed, W. et al. Comparison of virus concentration methods for the RT-qPCR-based recovery of murine hepatitis virus, a surrogate for SARS-CoV-2 from untreated wastewater. Sci. Total Environ. 739, 1–9 (2020).

22. Alpaslan-Kocamemi, B. et al. SARS-CoV-2 Detection in Istanbul Wastewater Treatment Plant Sludges. medRxiv (2020) doi:10.1101/2020.05.12.20099358.

23. Alpaslan Kocamemi, B. et al. First Data-Set on SARS-CoV-2 Detection for Istanbul Wastewaters in Turkey Authors Marmara University, Department of Environmental Engineering, Istanbul, Turkey Saglik Bilimleri University, Faculty of Medicine, Department of Medical Biology, Istanbul. *medRxiv* 1–10 (2020) doi:10.1101/2020.05.03.20089417.

24. Balboa, S. et al. THE FATE OF SARS-COV-2 IN WWTPS POINTS OUT THE SLUDGE LINE AS A SUITABLE SPOT FOR MONITORING. *medRxiv* 1–24 (2020).

25. Vandesompele, J. et al. Accurate normalization of real-time quantitative RT-PCR data by geometric averaging of multiple internal control genes. J. Artif. Intell. Res. 50, 1–30 (2002).

26. Riedel, G. et al. An extended ΔCT-method facilitating normalisation with multiple reference genes suited for quantitative RT-PCR analyses of human hepatocyte-like cells. PLoS One 9, 2–6 (2014).

27. Dawson, D. J., Paish, A., Staffell, L. M., Seymour, I. J. & Appleton, H. Survival of viruses on fresh produce, using MS2 as a surrogate for norovirus. J. Appl. Microbiol. 98, 203–209 (2005).

28. Colson, P. et al. Pepper mild mottle virus, a plant virus associated with specific immune responses, fever, abdominal pains, and pruritus in humans. PLoS One 5, (2010).

29. Ganzenmueller, T., Kluba, J., Becker, J. U., Bachmann, O. & Heim, A. Detection of cytomegalovirus (CMV) by real-time PCR in fecal samples for the non-invasive diagnosis of CMV intestinal disease. J. Clin. Virol. 61, 517–522 (2014).

30. Iyengar, V., Albaugh, G. P., Lohani, A. & Nair, P. P. Human stools as a source of viable colonic epithelial cells. FASEB J. (1991) doi:10.1096/fasebj.5.13.1655550.

31. Desilets, D. J. et al. Lectin Binding to Human Colonocytes Is Predictive of Colonic Neoplasia. Am. J. Gastroenterol. (1999) doi:10.1111/j.1572-0241.1999.00946.x.

32. Gies, A., Cuk, K., Schrotz-King, P. & Brenner, H. Fecal immunochemical test for hemoglobin in combination with fecal transferrin in colorectal cancer screening. United Eur. Gastroenterol. J. 6, 1223–1231 (2018).

33. Ottawa Public Health. City of Ottawa, Public Health Department Covid-19 Data Portal. https://www.ottawapublichealth.ca/en/reports-research-and-statistics/daily-covid19-dashboard.aspx (2020).

34. Kaplan, E. H. et al. Aligning SARS-CoV-2 Indicators via an Epidemic Model: Application to Hospital Admissions and RNA Detection in Sewage Sludge. *medRxiv* (2020) doi:10.1101/2020.06.27.20141739.

35. Cheung, K. S. et al. Gastrointestinal Manifestations of SARS-CoV-2 Infection and Virus Load in Fecal Samples From a Hong Kong Cohort: Systematic Review and Meta-analysis. Gastroenterology 159, 81–95 (2020).

36. Sultan, S. et al. AGA Institute Rapid Review of the Gastrointestinal and Liver Manifestations of COVID-19, Meta-Analysis of International Data, and Recommendations for the Consultative Management of Patients with COVID-19. Gastroenterology 159, 320-334.e27 (2020).

37. Zheng, S. et al. Viral load dynamics and disease severity in patients infected with SARS-CoV-2 in Zhejiang province, China, January-March 2020: Retrospective cohort study. BMJ 369, 1–8 (2020).

38. Wölfel, R. et al. Virological assessment of hospitalized patients with COVID-2019. Nature (2020) doi:10.1038/s41586-020-2196-x.

39. Chen, Y. et al. The presence of SARS-CoV-2 RNA in the feces of COVID-19 patients. J. Med. Virol. 92, 833–840 (2020).

40. He, X. et al. Temporal dynamics in viral shedding and transmissibility of COVID-19. Nat. Med. (2020) doi:10.1038/s41591-020-0869-5.

41. Bivins, A. COVID-19 Wastewater-Based Epidemiology: Methods and Insights for SARS-CoV-2 Community Surveillance. Genome webinar (2020).

42. Zaiontz, C. Multiple Correlation. http://www.real-statistics.com/correlation/multiple-correlation/.

43. Randazzo, W. et al. SARS-CoV-2 RNA in wastewater anticipated COVID-19 occurrence in a low prevalence area. Water Res. 181, (2020).

44. Randazzo, W., Cuevas-Ferrando, E., Sanjuán, R., Domingo-Calap, P. & Sánchez, G. Metropolitan Wastewater Analysis for COVID-19 Epidemiological Surveillance. *medRxiv* 1–8 (2020) doi:10.2139/ssrn.3586696.

45. Rosa, G. La et al. First detection of SARS-CoV-2 in untreated wastewaters in Italy. Sci. Total Environ. 736, 139652 (2020).

46. Peccia, J. et al. SARS-CoV-2 RNA concentrations in primary municipal sewage sludge as a leading indicator of COVID-19 outbreak dynamics. medRxiv 1, 2020.05.19.20105999 (2020).

